# Emergence and Rising Prevalence of Artemisinin Partial Resistance Marker Kelch13 P441L in a Low Malaria Transmission Setting in Southern Zambia

**DOI:** 10.1101/2025.01.02.24319706

**Authors:** Anne C. Martin, Jacob M. Sadler, Alfred Simkin, Michael Musonda, Ben Katowa, Japhet Matoba, Jessica Schue, Edgar Simulundu, Jeffrey A. Bailey, William J. Moss, Jonathan J. Juliano, Abebe A. Fola

## Abstract

Increasing artemisinin partial resistance (ArtR) due to mutations in the gene encoding Kelch13 (*Pfk13*) protein in eastern Africa is of urgent concern, and mutations, such as *Pfk13* P441L, continue to emerge. We used an amplicon deep-sequencing panel to estimate the prevalence of ArtR *Pfk13* mutations in samples collected between 2018 and 2023 in southern Zambia. *Pfk13* P441L was present in 30 of 501 samples (6%), and prevalence increased over time (0% to 7.2%). Further studies of the P441L mutation are needed to document its geographical origin, distribution and impact on treatment outcomes.

## Background

Malaria remains a critical public health problem in Africa, where the 246 million cases and 560,000 deaths in 2023 accounted for 94% of global cases and 95% of global deaths (1). Artemisinin-based combination therapies (ACTs) are vital for malaria control but their efficacy is jeopardized by emerging *Plasmodium falciparum* resistance. Artemisinin partial resistance (ArtR) due to Kelch13 (*Pfk13*) propeller-domain mutations was first detected in Cambodia in 2006 and quickly spread across Southeast Asia (2). In Africa, *Pfk13*-associated ArtR was first identified in Rwanda (*Pfk13* R561H) in 2014 (3), followed by Uganda (A675V, C469Y) in 2016 (4), and Ethiopia and Eritrea (R622I) also in 2016 (5).

These mutations are increasing in prevalence and geographic range. Countries across East Africa and the Horn of Africa (Eritrea, Ethiopia, Uganda, Rwanda) have recorded a high prevalence of *Pfk13* mutations across multiple sites, and the neighboring countries of Kenya and Zambia recently detected these mutations at low prevalence (6–10). Despite the increase in *Pfk13* mutations associated with ArtR in the East and Horn of Africa, there are not yet reports of *Pfk13* mutations in Central and Southern Africa at a prevalence greater than the World Health Organization (WHO) 5% threshold to signal concern.

P441L is a highly prevalent mutation in Myanmar (11), first appearing in Africa in 2018 in single samples from north-western Zambia (10) and Rwanda (3). P441L is considered a candidate marker of resistance by the WHO and has been associated with delayed parasite clearance in Southeast Asia in addition to its high prevalence in Myanmar (12). More recent surveys have detected P441L at sites in multiple countries in Africa, reaching a prevalence of 12 to 23% in parts of Uganda (7), respective frequencies of 7.7% and 17.2% at sites in Rwanda and Tanzania (8), in 4 samples from Ethiopia (9), and in one sample in Democratic Republic of Congo (DRC) (16).

Zambia has regions of low malaria transmission, considered favorable environments for accelerating the emergence of antimalarial drug resistance. With smaller parasite populations and the relative absence of host premunition, symptomatic infections have high parasite biomass, fewer competing clones, and are subject to high treatment rates, all of which allow mutant, otherwise-less-fit, resistant parasites to better compete with the wildtype. Thus, although the relative scarcity of malaria cases in low-transmission regions undermines efficiency in gathering sufficient samples for molecular surveillance, these are the key areas to survey for foci of emerging drug resistance. This study used amplicon deep-sequencing to estimate the prevalence and frequency of markers of ArtR and partner drug resistance in samples collected across a six-year period in a low transmission area in southern Zambia.

## Methods

The study site included the catchment areas of four health centers in Choma District, Southern Province, Zambia where the 5-year average malaria incidence was 10 cases per 1000 person-years. Dried blood spots (DBS) were collected on Whatman 3MM chromatography paper through health-center and community surveillance from 2018 to 2023. Health-center surveillance enrolled and sampled symptomatic individuals with a positive rapid diagnostic test (RDT) from two health centers in 2018 - 2021 and from four health centers in 2022 - 2023. Community surveillance occurred through two successive field studies, a 200-household cohort in 2018 - 2021 and a malaria focal hotspot study from 2022 - 2023. Both studies enrolled and sampled entire households irrespective of individual malaria infection status, with their samples included if they were determined to be parasitemic using quantitative polymerase chain reaction (qPCR) targeting the multicopy *P. falciparum* mitochondrial cytochrome b (*Pfcytb)* gene. Details on study design, sampling, and enrollment and eligibility criteria can be found in **Table S1**.

All samples were further screened using qPCR targeting the lactate dehydrogenase (*Pfldh)* gene to quantify parasitemia with a single copy gene and determine further sample processing. Quantification of parasitemia was done using five standard curves of one to ten thousand parasites per microliter (µL) (**Figure S1**). All samples positive for *Pfldh* and samples negative for *Pfldh* but positive in duplicate for *Pfcytb* were included. Selective whole genome amplification (sWGA) was performed on all samples (n=183) with fewer than 50 parasites/µL using a published protocol (13). Samples confirmed to contain *P. falciparum* DNA were genotyped in duplicate using *Pf-SMARRT*, a 24-locus deep sequencing amplicon panel that targets key markers on drug resistance genes, including multidrug-resistance 1 (*Pfmdr1*), chloroquine resistance transporter (*Pfcrt*), dihydrofolate reductase (*Pfdhfr*), dihydropteroate synthase (*Pfdhps*), and 21 validated and candidate mutations in *Pfk13* (14). Sequenced data demultiplexing, quality control, mapping against the reference genome, and variant calling were done using SeekDeep software, with a minimum of 100 reads per replicate and minimum of 1% read frequency to call haplotypes (https://github.com/bailey-lab/SeekDeep). All loci were checked across replicate pairs for agreement in haplotype count and amino acid frequency. All downstream analysis and data visualization were performed using R (version 4.2.3). Population level prevalence of drug resistance was defined as the proportion of samples successfully sequenced at the given loci that contained the mutated allele at any within-sample frequency. Allele population frequency was defined as the proportion of all sequenced reads in the population that contained that mutation. Sulfadoxine-pyrimethamine (SP) haplotypes were inferred from population level allele frequencies of SP resistance markers.

## Results

The assay was run on 570 samples and 540 samples were successfully sequenced at one or more loci to determine the annual prevalence of drug resistance markers (**Table 1**). sWGA was done for 183 of these samples and 130 samples were successfully sequenced at one or more loci. The supplement contains data on annual allele frequencies (**Table S2**), demographic characteristics of sampled individuals (**Tables S3, S4**), and parasitemia of included samples (**Figure S1**).

**Table 1.** Prevalence of mutations by year for markers or potential markers of drug resistance.

| Gene | Mutation | All |  | 2018 |  | 2019 |  | 2020 |  | 2021 |  | 2022 |  | 2023 |  |
| --- | --- | --- | --- | --- | --- | --- | --- | --- | --- | --- | --- | --- | --- | --- | --- |
|  |  | n† | Prev*. (%) | n | Prev. (%) | n | Prev. (%) | n | Prev. (%) | n | Prev. (%) | n | Prev. (%) | n | Prev. (%) |
| *Pfdhfr* | S108N | 480 | 99.6 | 7 | 100 | 47 | 95.7 | 122 | 100 | 18 | 100 | 55 | 100 | 209 | 100 |
| *Pfdhfr* | N51I | 511 | 97.1 | 8 | 100 | 52 | 86.5 | 140 | 97.9 | 19 | 94.7 | 57 | 100 | 214 | 98.1 |
| *Pfdhfr* | C59R | 511 | 95.1 | 8 | 100 | 52 | 88.5 | 140 | 98.6 | 19 | 100 | 57 | 93 | 214 | 95.3 |
| *Pfdhps* | A437G | 479 | 94.2 | 7 | 57.1 | 48 | 89.6 | 130 | 96.2 | 17 | 88.2 | 53 | 98.1 | 203 | 95.1 |
| *Pfdhps* | K540E | 334 | 86.5 | 4 | 25 | 30 | 80 | 92 | 84.8 | 13 | 92.3 | 30 | 86.7 | 149 | 89.9 |
| *Pfdhps* | A581G | 467 | 3.6 | 7 | 0 | 40 | 2.5 | 122 | 1.6 | 19 | 5.3 | 52 | 1.9 | 207 | 5.8 |
| *Pfk13* | P441L | 501 | 6 | 9 | 0 | 50 | 6 | 140 | 3.6 | 19 | 5.3 | 54 | 9.3 | 209 | 7.2 |
| *Pfk13* | A578S | 513 | 0.6 | 9 | 0 | 52 | 0 | 146 | 0 | 20 | 0 | 54 | 0 | 211 | 1.4 |
| *Pfmdr1* | Y184F | 508 | 41.3 | 8 | 50 | 52 | 57.7 | 144 | 30.6 | 20 | 35 | 56 | 39.3 | 207 | 44.9 |
| *Pfmdr1* | N86 | 459 | 100 | 7 | 100 | 43 | 100 | 114 | 100 | 18 | 100 | 51 | 100 | 205 | 100 |

\*Mutant samples include samples with presence of the drug resistant allele at any frequency
†n= number of samples successfully genotyped

### Rising prevalence of the candidate Pfk13 ArtR marker P441L

We assessed the prevalence of WHO-validated or candidate *Pfk13* mutations associated with ArtR. The WHO-candidate *Pfk13* P441L mutation was present in 6% (30 of 501) of all samples and prevalence increased over the study period, from 0% in 2018 to 7.2% in 2023, with a high of 9.3% in 2022 (**Figure 1, Table 1**). P441L mutants were found at all four health centers and in all years except 2018. The age and sex distributions did not differ across individuals with and without samples with P441L (**Table S4**). Notably, 22% of individuals with P441L reported travel in the last four weeks compared to 16% without the mutation. Individuals harboring parasites with P441L mutations reported travel to Monze District in Southern Province as well as Central and Northwestern Provinces. No individuals reported international travel. In addition to the P441L mutation, the *Pfk13* A578S mutation was identified in parasites from 3 individuals (0.6%) sampled in 2023.

**Figure 1.**
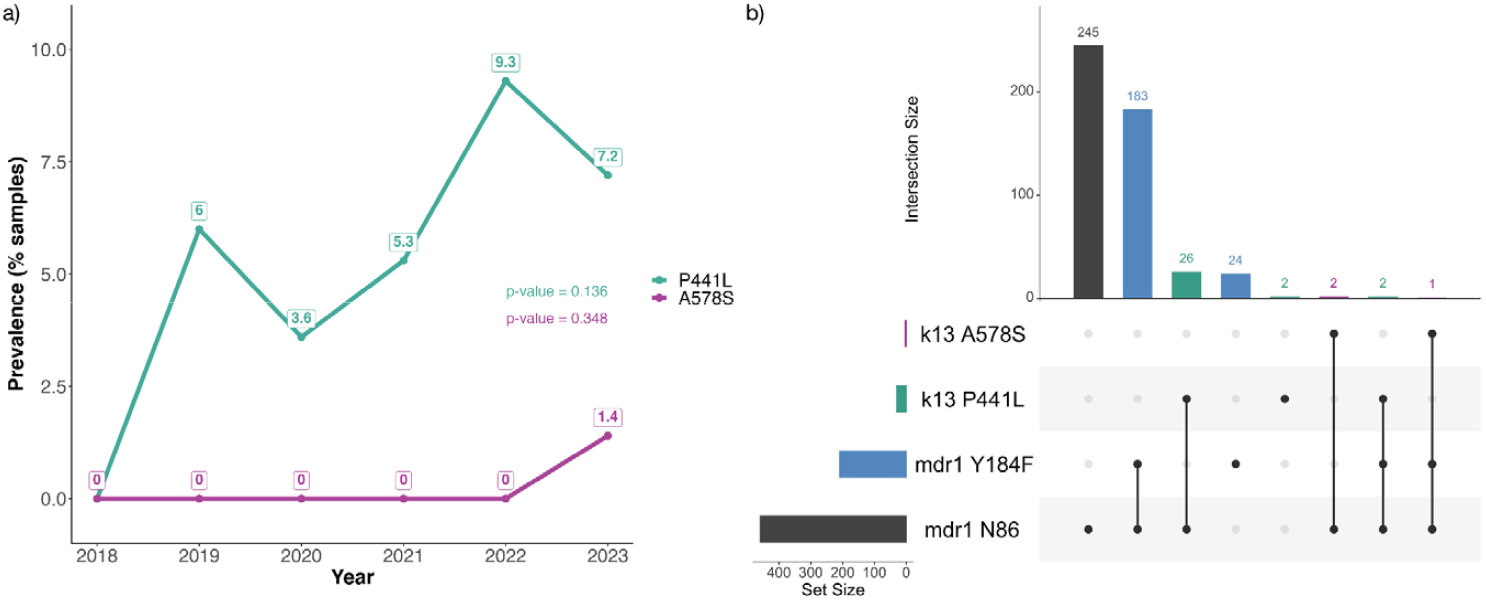
Kelch13 (*Pfk13*) mutation P441L has been detected in Southern Zambia. a) Prevalence of P441L was 0% in 2018, reached a high of 9.3% in 2022, and was 7.2% in 2023. Prevalence of *Pfk13* mutation A578S was 0% all years until 2023 when prevalence was 1.4%. Jonckheere’s trend test was used to evaluate temporal changes in prevalence and generate p-values. b) The upset plot for *Pfk13* mutations and *Pfmdr1* mutations shows the number of samples containing each mutation (horizontal bars) and the number of combinations of mutations found within the same sample (vertical bars). There were two samples where *Pfk13* P441L and *Pfmdr1* Y184F co-occurred.

### Markers of artemisinin partner drug and sulfadoxine-pyrimethamine resistance

The overall prevalence of MDR1 Y184F, which is associated with chloroquine and amodiaquine resistance, was 41%, and there was no change in prevalence over time (**Table 1, Figure S2**). The wildtype MDR1 N86 is associated with possible reduced susceptibility to lumefantrine and was present in all samples. There were two samples in which *Pfk13* P441L and *Pfmdr1* Y184F mutations co-occurred (**Figure 1**). As expected, the prevalence of mutations in resistance markers to SP was high (**Figures S3, S4**).

## Discussion

The expanded presence of antimalarial drug resistance markers beyond East Africa and the Horn of Africa, now to southern Africa, is a growing threat to malaria control. We used an amplicon panel to examine the prevalence of genetic markers associated with drug resistance at a low transmission site in southern Zambia. Two mutations within the *Pfk13* propeller domain were present at low frequencies – P441L first appearing in 2019 and A578S in 2023. No other previously reported, non-synonymous mutations in *Pfk13* were detected. While A578S is found across the continent, this mutation is not associated with artemisinin resistance (15). On the other hand, P441L is associated with delayed clearance in Southeast Asia and is at high prevalence in Myanmar despite competition with other validated mutations. Therefore, P441L’s emergence in several countries in Africa, where its prevalence is similarly outpacing that of other common validated mutations, is cause for significant concern. In southern Zambia the prevalence of P441L in 2023 was 7.2% and the six-year prevalence was 6%. Both exceed the 5% threshold used by the WHO to indicate a concerning level of resistance warranting further investigation and monitoring. The predominance of MDR1 N86 (wildtype) heightens this concern, as the combination of artemether and lumefantrine is first-line treatment in Zambia.

The widespread emergence of the P441L mutation spanning Ethiopia, Tanzania, Rwanda, Uganda, and now the DRC and Zambia has occurred without a clearly identifiable epicenter. This pattern does not fit with a simple emergence-and-spread model typically observed for artemisinin resistance (ArtR) mutations. If resistance mutations spread in a straightforward manner, one might expect highly prevalent and wider distributed ArtR mutations such as 561H, 675V, 459F/Y, or 622I to first appear in Zambia.

Instead, the singular detection of P441L in Zambia raises the possibility of an undiscovered region with high P441L prevalence and limited surveillance, potentially the source for its seeding across multiple sites and nations. Alternatively, the rise of P441L could reflect multiple independent emergences. The increasing prevalence of P441L in Southeast Asia, also rising quickly relative to that of earlier arising mutations, has been linked to partner drug or compensatory mutations elsewhere in the genome (11). The conspicuous emergence and persistence of P441L in multiple African countries could therefore be driven, at least in part, by genomic interactions and other biological factors that confer advantages to this mutation. For example, in Rwanda, where P441L co-occurs with other *Pfk13* resistance markers, understanding whether these interactions are synergistic, compensatory, or coincidental could provide insights into its persistence and fitness (7). Also, while P441L only co-occurred with *Pfmdr1* Y184F in two samples, it could be associated with evolving partner drug resistance, which was thought to drive the *Pfk13* C580Y mutation to high frequency in Southeast Asia.

There are several limitations to this work. First, there is moderate missingness in loci, particularly in the low parasitemia samples, and therefore we do not have complete drug resistance profiles. *Pf-SMARRT* does not cover the entire *Pfk13* propeller domain, although it includes all but one (amino acid site 574) of the WHO-candidate and validated mutations (14). Another important limitation is the sample size variation due to the geographic scale and sampling of the parent studies. Smaller sample size reduced the precision of the prevalence estimates and the sensitivity to detect emergent mutations. Emergence of P441L may have happened earlier than 2019. The 2022 expansion to two additional health centers increased sample size, but is unlikely to have introduced temporal bias, as the geographic distribution of the P441L mutation was proportional to the geographic distribution of all infections (**Table S4**). The change in community surveillance from sampling a continuous cohort to sampling communities of index malaria cases will bias estimates of parasitemia prevalence but not estimates of mutation prevalence, since inclusion in this analysis was conditional on parasitemia. Overall, the longitudinal surveillance across these multiple studies, and the ability to sample and sequence both symptomatic and low-density asymptomatic infections, are strengths of this work.

The P441L mutation is present and likely increasing in prevalence in *P. falciparum* in southern Zambia, consistent with a hypothesis that this loci is under selective pressure. Still, there are gaps in our knowledge of the implications of this mutation for malaria treatment. First, we do not understand the extent to which the mutation is associated with ArtR. Therapeutic efficacy studies with a focus on P441L and further ex-vivo studies are needed. Second, the unexpected rise in prevalence of P441L suggests possible genome-level interactions and/or biological advantages driving selection mechanisms. Whole genome sequencing of samples containing P441L should be done to identify associated mutations and provide a comprehensive understanding of the genetic landscape and insights into underlying mechanisms. Third, the increase in prevalence and geographic distribution of *Pfk13* mutations in Africa is alarming and should serve as a call to embed molecular surveillance for emergent drug resistance mutations into routine national and multinational surveillance.

## Ethics

The samples and data used in this study were a part of two IRB-approved studies. The “Against Transmission Of Malaria With Everyone (ANTOOMWE) study” had ethical approval from the Johns Hopkins Bloomberg School of Public Health (Baltimore, Maryland) under IRB no: 00003467 and the Tropical Diseases Research Center (TDRC under IRB no: TDRC/ERC/2010/14/11. The “Magnifying the Utility of Surveillance in Elimination-focused Malaria Operations (MUSEMO)” study has ethical approval from the Johns Hopkins Bloomberg School of Public Health (Baltimore, Maryland) under IRB no: 00019447 and Macha Research Trust (Macha, Zambia) under IRB no: 0007649. Written informed consent was obtained from adults 18 years and older, and parental permission from parents or guardians of individuals younger than 18 years. Oral assent was obtained from children between 12 and 18 years.

## Data availability

Sequencing data are available at (SRA number pending). Under the National Health Research Act, the Government of Zambia does not allow public access to metadata collected in Zambia. All investigators interested in the datasets supporting the conclusions of this article are required to submit a written request to the Ministry of Health. Contact the Macha Research Trust IRB Chairperson (, +260979402560) to coordinate the request. Analytic code for the analysis can be found here: https://github.com/anniecmartin/ (to be posted after revisions)

## Funding

This work was supported by funds from the National Institute of Allergy and Infectious Diseases (NIAID) of the National Institutes of Health [grant numbers U19AI089680 to WJM, K24AI134990 to JJJ and T32AI138953-03 to AM], the Bloomberg Philanthropies, and the Johns Hopkins Malaria Research Institute.

## Supplementary data

Supplementary data are available at JID online.

## Author contributions

ACM oversaw data collection and management for the MUSEMO study, conducted epidemiological analysis, and drafted the manuscript. JMS prepared and ran the amplicon panel on all samples. JMS and AS ran the bioinformatics pipeline processing of sequencing data. MM and BK conducted lab assays for identifying samples for inclusion. JM managed data collection, cleaning, and sample organizing across both studies. JS oversaw data collection and management for the ANTOOMWE study. AAF, JAB, and JJJ led conceptualization of the manuscript and along with ES and WJM, contributed to the drafts. All authors contributed to manuscript revisions and provided final approval.

## Acknowledgements

The following reagent was obtained through BEI Resources, NIAID, NIH: *Plasmodium falciparum*, Strain 3D7, MRA-102, contributed by Daniel J. Carucci. The following reagent was obtained through BEI Resources, NIAID, NIH: Genomic DNA from *Plasmodium falciparum*, Strain 3D7, MRA-102G, contributed by Daniel J. Carucci. We thank the Macha Research Trust study team members, health center staff, and community health workers who collected samples and data. We thank the National Malaria Elimination Centre, Southern Province and Choma District leadership for their support. Lastly, we thank all of the MUSEMO and ANTOOMWE study participants for their participation and time.

## Conflicts of interest

Authors have no competing interests to declare.

## Prior presentation

This work has not been presented elsewhere.

## Notes

### Competing Interest Statement

The authors have declared no competing interest.

### Author Declarations

The samples and data used in this study were a part of two IRB-approved studies. The Against Transmission Of Malaria With Everyone (ANTOOMWE) study had ethical approval from the Johns Hopkins Bloomberg School of Public Health (Baltimore, Maryland) under IRB no: 00003467 and the Tropical Diseases Research Center (TDRC under IRB no: TDRC/ERC/2010/14/11. The Magnifying the Utility of Surveillance in Elimination-focused Malaria Operations (MUSEMO) study has ethical approval from the Johns Hopkins Bloomberg School of Public Health (Baltimore, Maryland) under IRB no: 00019447 and Macha Research Trust (Macha, Zambia) under IRB no: 0007649. Written informed consent was obtained from adults 18 years and older, and parental permission from parents or guardians of individuals younger than 18 years. Oral assent was obtained from children between 12 and 18 years.

