## Supplement for "Emergence and Rising Prevalence of Artemisinin Partial Resistance Marker Kelch13 P441L in a Low Malaria Transmission Setting in Southern Zambia"

**Brief Report**

**Table S1. Surveillance, study design, and sampling strategies of parent studies.** The 570 samples analyzed were collected through passive surveillance at health centers and active surveillance through two parent studies. This table describes the study area, study design, and sampling frequency for each of the studies and surveillance methods across the years. We provide the total samples positive using quantitative polymerase chain reaction (qPCR) for *P. falciparum* mitochondrial cytochrome b (*Pfcytb)* gene, the subset of those that were also positive for lactate dehydrogenase (*Pfldh)*, the total samples selected to run on the assay, and the subset of those for which selective whole genome amplification (sWGA) was performed.

| **Surveillance Type and Location** | **Parent study** | **Years** | **Study area** | **Study design** | **Eligibility** | **Sampling frequency** | **Total qPCR positives** | | **Total (sWGA) samples run** | **Total**  **(sWGA) samples successful** |
| --- | --- | --- | --- | --- | --- | --- | --- | --- | --- | --- |
|  |  |  |  |  |  |  | **cytb** | **LDH** |  |  |
| Passive, at health-center | ANTOOMWE | 2018  2019  2020  2021 | Macha Hospital;  Mapanza Rural Health Centre | Health-center case study | 6+ month-old, symptomatic, RDT positives | Once, at time of diagnosis | 235 | 226 | 226 (42) | 226 (42) |
|  | MUSEMO | 2022  2023 | Above, plus:  Mbabala Rural Health Centre;  Mangunza Rural Health Centre |  |  |  | 302 | 279 | 290 (58) | 284 (51) |
| Active, in community | ANTOOMWE | 2018  2019  2020  2021 | Mapanza Rural Health Centre | 200-household cohort | 6+ month-old, cohort household members | Monthly, for 24 months | 144 | 11 | 41 (37) | 11 (7) |
|  | MUSEMO | 2022  2023 | Macha Hospital  Mapanza Rural Health Centre | Reactive surveillance in index case communities | 6+ month-old, community members living within 250 m of index case | Twice, 7 and 35 days after index diagnosis | 78 | 21 | 53 (46) | 46 (30) |

Abbreviations: Against Transmission Of Malaria With Everyone (ANTOOMWE); [Magnifying the Utility of Surveillance in Elimination-focused Malaria Operations](https://phirst.jhsph.edu/sph/sd/Rooms/DisplayPages/LayoutInitial?Container=com.webridge.entity.Entity%5bOID%5bC4CC710EBE4D0F4FA4FA8080268C6905%5d%5d) (MUSEMO), quantitative polymerase chain reaction (qPCR); *P. falciparum* mitochondrial cytochrome b (*cytb)*; lactate dehydrogenase (*ldh)*; selective whole genome amplification (sWGA).

**Table S2a. Annual prevalence of validated or potential markers of antimalarial drug resistance.**

|  |  | **2018** | | | **2019** | | | **2020** | | | **2021** | | | **2022** | | | **2023** | | |
| --- | --- | --- | --- | --- | --- | --- | --- | --- | --- | --- | --- | --- | --- | --- | --- | --- | --- | --- | --- |
| **Gene** | **Mutation** | **n** | **Prev. (%)** | **95% CI** | **n** | **Prev. (%)** | **95% CI** | **n** | **Prev. (%)** | **95% CI** | **n** | **Prev. (%)** | **95% CI** | **n** | **Prev. (%)** | **95% CI** | **n** | **Prev. (%)** | **95% CI** |
| ***Pfdhfr*** | S108N | 7 | 100 | 100, 100 | 47 | 95.7 | 89.9, 100 | 122 | 100 | 100, 100 | 18 | 100 | 100, 100 | 55 | 100 | 100, 100 | 209 | 100 | 100, 100 |
| ***Pfdhfr*** | N51I | 8 | 100 | 100, 100 | 52 | 86.5 | 77.2, 95.8 | 140 | 97.9 | 95.5, 100 | 19 | 94.7 | 84.6, 100 | 57 | 100 | 100, 100 | 214 | 98.1 | 96.3, 99.9 |
| ***Pfdhfr*** | C59R | 8 | 100 | 100, 100 | 52 | 88.5 | 79.8, 97.2 | 140 | 98.6 | 96.7, 100 | 19 | 100 | 100, 100 | 57 | 93 | 86.4, 99.6 | 214 | 95.3 | 92.5, 98.1 |
| ***Pfdhps*** | A437G | 7 | 57.1 | 20.4, 93.8 | 48 | 89.6 | 81, 98.2 | 130 | 96.2 | 92.9, 99.5 | 17 | 88.2 | 72.9, 100 | 53 | 98.1 | 94.4, 100 | 203 | 95.1 | 92.1, 98.1 |
| ***Pfdhps*** | K540E | 4 | 25 | 0, 67.4 | 30 | 80 | 65.7, 94.3 | 92 | 84.8 | 77.5, 92.1 | 13 | 92.3 | 77.8, 100 | 30 | 86.7 | 74.5, 98.9 | 149 | 89.9 | 85.1, 94.7 |
| ***Pfdhps*** | A581G | 7 | 0 | 0, 0 | 40 | 2.5 | 0, 7.3 | 122 | 1.6 | 0, 3.8 | 19 | 5.3 | 0, 15.4 | 52 | 1.9 | 0, 5.6 | 207 | 5.8 | 2.6, 9 |
| ***Pfk13*** | P441L | 9 | 0 | 0, 0 | 50 | 6 | 0, 12.6 | 140 | 3.6 | 0.5, 6.7 | 19 | 5.3 | 0, 15.4 | 54 | 9.3 | 1.6, 17 | 209 | 7.2 | 3.7, 10.7 |
| ***Pfk13*** | A578S | 9 | 0 | 0, 0 | 52 | 0 | 0, 0 | 146 | 0 | 0, 0 | 20 | 0 | 0, 0 | 54 | 0 | 0, 0 | 211 | 1.4 | 0, 3 |
| ***Pfmdr1*** | Y184F | 8 | 50 | 15.4, 84.6 | 52 | 57.7 | 44.3, 71.1 | 144 | 30.6 | 23.1, 38.1 | 20 | 35 | 14.1, 55.9 | 56 | 39.3 | 26.5, 52.1 | 207 | 44.9 | 38.1, 51.7 |
| ***Pfmdr1*** | N86 | 7 | 100 | 100, 100 | 43 | 100 | 100, 100 | 114 | 100 | 100, 100 | 18 | 100 | 100, 100 | 51 | 100 | 100, 100 | 205 | 100 | 100, 100 |

**Table S2b. Annual frequency of validated or potential markers of antimalarial drug resistance.** Allele population frequency was defined as the proportion of reads containing that mutation across all sequenced reads in the population. Confidence intervals were estimated using the Wald interval for proportions.

|  |  | **2018** | | | **2019** | | | **2020** | | | **2021** | | | **2022** | | | **2023** | | |
| --- | --- | --- | --- | --- | --- | --- | --- | --- | --- | --- | --- | --- | --- | --- | --- | --- | --- | --- | --- |
| **Gene** | **Mutation** | **Read count** | **Freq. (%)** | **95% CI** | **Read count** | **Freq. (%)** | **95% CI** | **Read count** | **Freq. (%)** | **95% CI** | **Read count** | **Freq. (%)** | **95% CI** | **Read count** | **Freq. (%)** | **95% CI** | **Read count** | **Freq. (%)** | **95% CI** |
| ***Pfdhfr*** | S108N | 38261 | 100 | 100, 100 | 293944 | 94 | 93.9, 94.1 | 633330 | 99.9 | 99.9, 99.9 | 57919 | 100 | 100, 100 | 344453 | 100 | 100, 100 | 2257412 | 100 | 100, 100 |
| ***Pfdhfr*** | N51I | 24307 | 100 | 100, 100 | 120649 | 93.3 | 93.2, 93.4 | 313680 | 99.9 | 99.9, 99.9 | 24394 | 96 | 95.8, 96.2 | 198146 | 99.8 | 99.8, 99.8 | 1772258 | 98 | 98, 98 |
| ***Pfdhfr*** | C59R | 24307 | 100 | 100, 100 | 120649 | 77 | 76.8, 77.2 | 313680 | 97.2 | 97.1, 97.3 | 24394 | 98.1 | 97.9, 98.3 | 198146 | 99 | 99, 99 | 1772258 | 94.5 | 94.5, 94.5 |
| ***Pfdhps*** | A437G | 10340 | 88.4 | 87.8, 89 | 157660 | 82.8 | 82.6, 83 | 488363 | 91.6 | 91.5, 91.7 | 23344 | 96.3 | 96.1, 96.5 | 143424 | 94.5 | 94.4, 94.6 | 1319716 | 97 | 97, 97 |
| ***Pfdhps*** | K540E | 1863 | 0 | 0, 0 | 32535 | 58.4 | 57.9, 58.9 | 112721 | 78.4 | 78.2, 78.6 | 4985 | 100 | 100, 100 | 43753 | 86.1 | 85.8, 86.4 | 630527 | 91.3 | 91.2, 91.4 |
| ***Pfdhps*** | A581G | 2743 | 0 | 0, 0 | 73025 | 0.3 | 0.3, 0.3 | 425765 | 2 | 2, 2 | 18784 | 12.2 | 11.7, 12.7 | 173965 | 2.6 | 2.5, 2.7 | 1325936 | 5.6 | 5.6, 5.6 |
| ***Pfk13*** | P441L | 6580 | 0 | 0, 0 | 143621 | 3.2 | 3.1, 3.3 | 392294 | 1.6 | 1.6, 1.6 | 17502 | 1.4 | 1.2, 1.6 | 115585 | 7 | 6.9, 7.1 | 883801 | 8.3 | 8.2, 8.4 |
| ***Pfk13*** | A578S | 6526 | 0 | 0, 0 | 166216 | 0 | 0, 0 | 441753 | 0 | 0, 0 | 24888 | 0 | 0, 0 | 139818 | 0 | 0, 0 | 1544835 | 0.9 | 0.9, 0.9 |
| ***Pfmdr1*** | Y184F | 17083 | 75.5 | 74.9, 76.1 | 177288 | 49.1 | 48.9, 49.3 | 314801 | 27 | 26.8, 27.2 | 26529 | 13.2 | 12.8, 13.6 | 160499 | 16 | 15.8, 16.2 | 1661263 | 43.5 | 43.4, 43.6 |
| ***Pfmdr1*** | N86 | 26011 | 100 | 100, 100 | 151282 | 100 | 100, 100 | 354620 | 100 | 100, 100 | 31958 | 100 | 100, 100 | 207706 | 100 | 100, 100 | 1269568 | 100 | 100, 100 |

**Table S3. Characteristics of participants for all *Plasmodium falciparum* infections and for which samples were successfully sequenced at one or more drug resistant loci.**

|  | All infections (n=570) | All sequenced (n = 540) |
| --- | --- | --- |
| Parasites/µL (median [IQR]) | 939 [125, 3416] | 1027 [148, 3616] |
| Facility diagnosed (%) | 502 (90)) | 493 (93) |
| Sex = Female (mean (SD)) | 0.45 (0.50) | 0.44 (0.50) |
| Age (y) (median [IQR]) | 16 [10, 25] | 16 [10, 24] |
| Year (%) |  |  |
| 2018 | 9 (1.6) | 8 (1.5) |
| 2019 | 57 (10.3) | 56 (10.7) |
| 2020 | 149 (27.0) | 146 (27.9) |
| 2021 | 22 ( 4.0) | 22 ( 4.2) |
| 2022 | 66 (12.0) | 63 (12.0) |
| 2023 | 249 (45.1) | 229 (43.7) |
| Travelled in last month (%) | 87 (15.2) | 80 (15.7) |

**Table S4. Characteristics of participants with and without P441L mutations.**

|  | P441 (n=510) | P441L (n=30) | p* |
| --- | --- | --- | --- |
| Parasites/µL median [IQR] | 1013 [144, 3664] | 1327 [408, 2904] | 0.453 |
| Facility diagnosed (%) | 465 (93.2) | 28 (93.3) | 1 |
| Health Centre (%) |  |  | 0.543 |
| Macha Hospital | 82 (17.3) | 4 (13.8) |  |
| Mangunza Rural Health Centre | 37 ( 7.8) | 1 (3.4) |  |
| Mapanza Rural Health Centre | 282 (59.4) | 17 (58.6) |  |
| Mbabala Rural Health Centre | 74 (15.6) | 7 (24.1) |  |
| Sex = Female (%) | 212 (44.9)) | 10 (34.5) | 0.365 |
| Age (y) median [IQR] | 16 [10, 25] | 16 [12, 20] | 0.811 |
| Year (%) |  |  | 0.693 |
| 2018 | 8 (1.6) | 0 (0.0) |  |
| 2019 | 52 (10.5) | 4 (13.3) |  |
| 2020 | 141 (28.5) | 5 (16.7) |  |
| 2021 | 21 ( 4.3) | 1 ( 3.3) |  |
| 2022 | 58 (11.7) | 5 (16.7) |  |
| 2023 | 214 (43.3) | 15 (50.0) |  |
| Travelled in last month = 1 (%) | 71 (15.6) | 6 (22.2) | 0.518 |
| *p values are derived from chi-squared tests for categorical variables and t-tests for continuous variables. | | | |

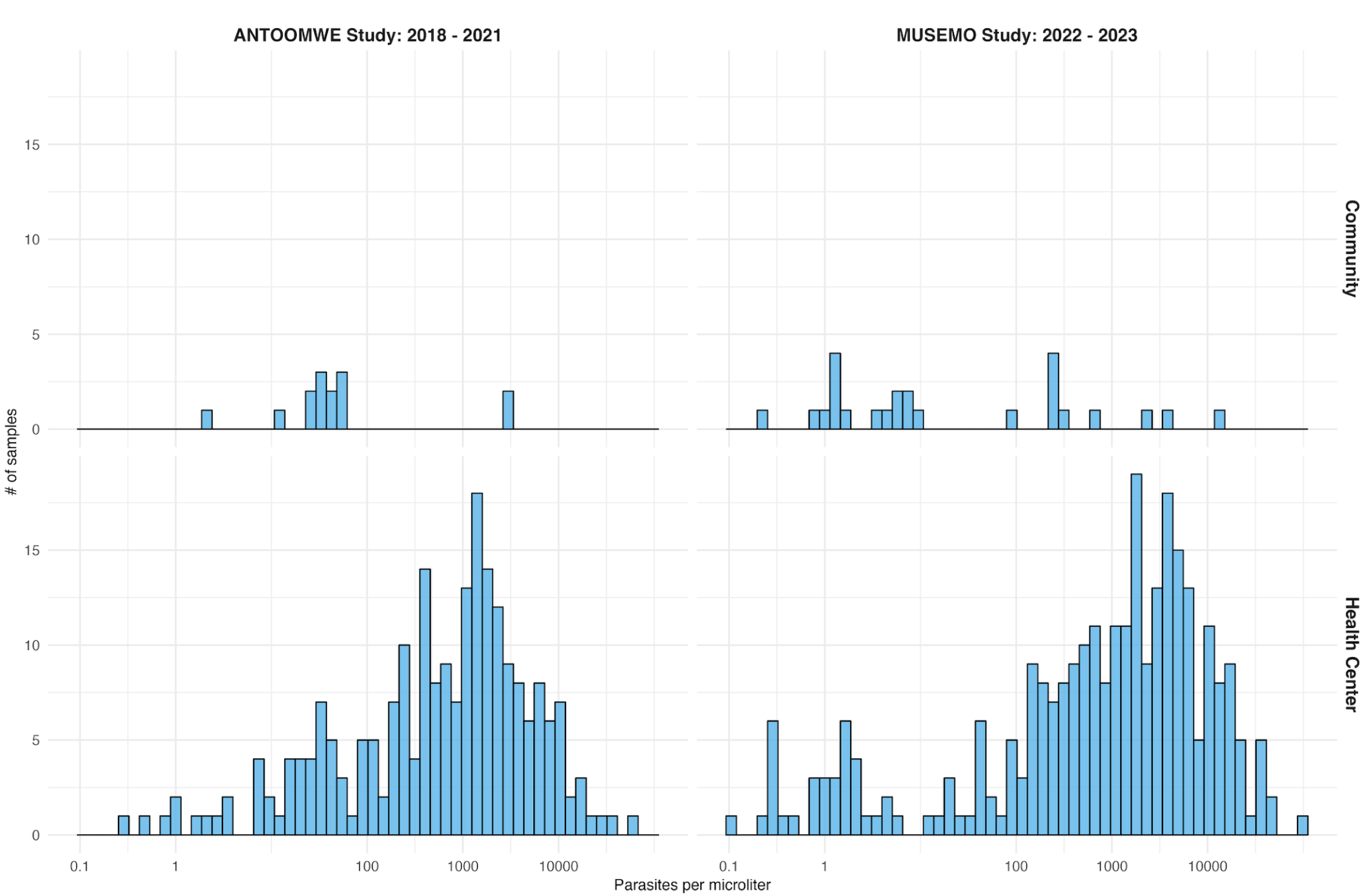

**Figure S1. Parasitemia from qPCR targeting the single copy *P. falciparum* lactate dehydrogenase (*Pfldh)* gene for each sample by study and surveillance method.** The median parasite density of successfully run samples was 1027 parasites/µL (full range 0.1 to 97719 parasites/μL). Abbreviations: Against Transmission Of Malaria With Everyone (ANTOOMWE); [Magnifying the Utility of Surveillance in Elimination-focused Malaria Operations](https://phirst.jhsph.edu/sph/sd/Rooms/DisplayPages/LayoutInitial?Container=com.webridge.entity.Entity%5bOID%5bC4CC710EBE4D0F4FA4FA8080268C6905%5d%5d) (MUSEMO)

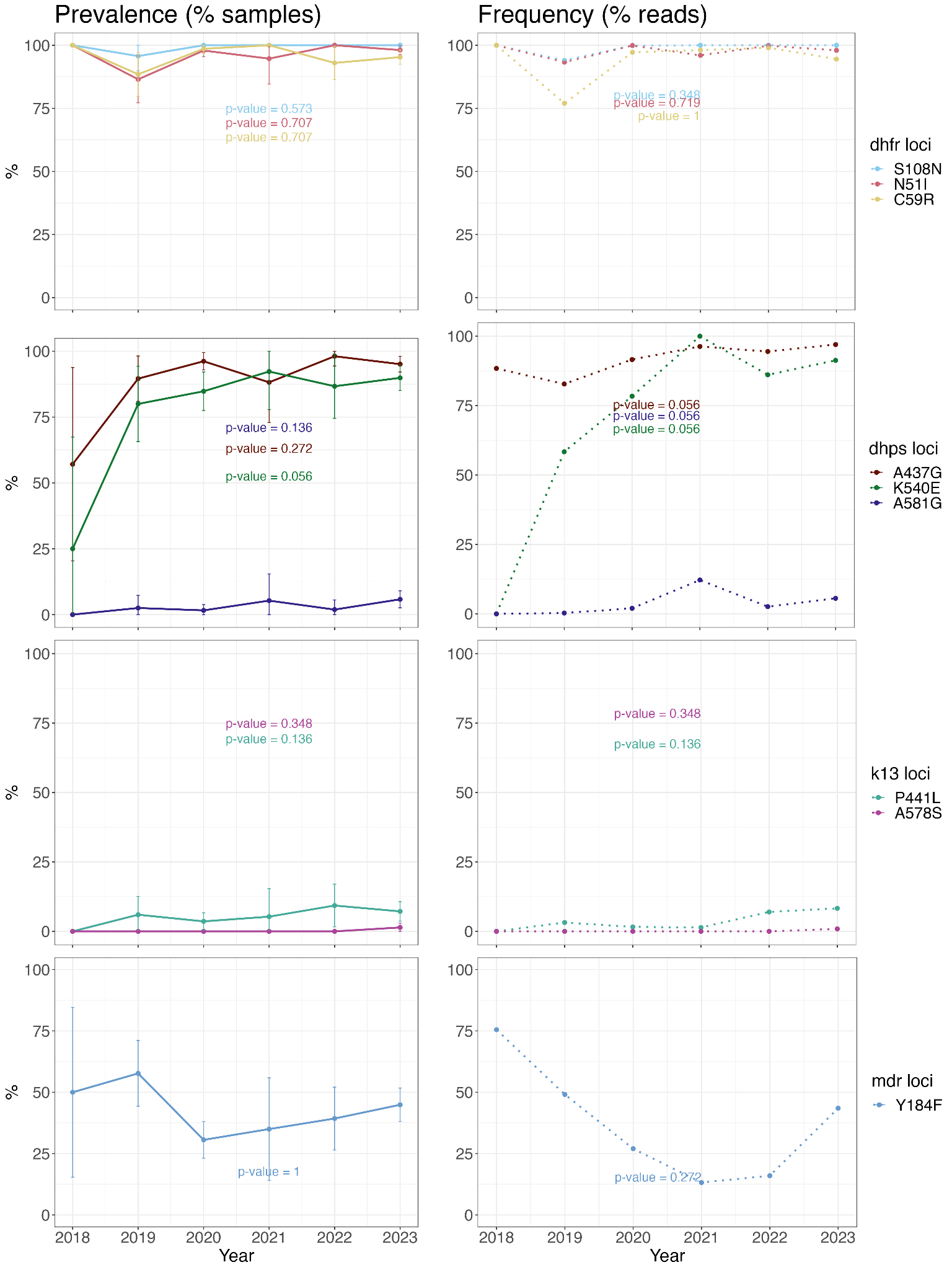

**Figure S2. Annual prevalence and frequency of mutations.** Jonckheere’s trend test was used to evaluate temporal changes in prevalence and frequency (p-values).

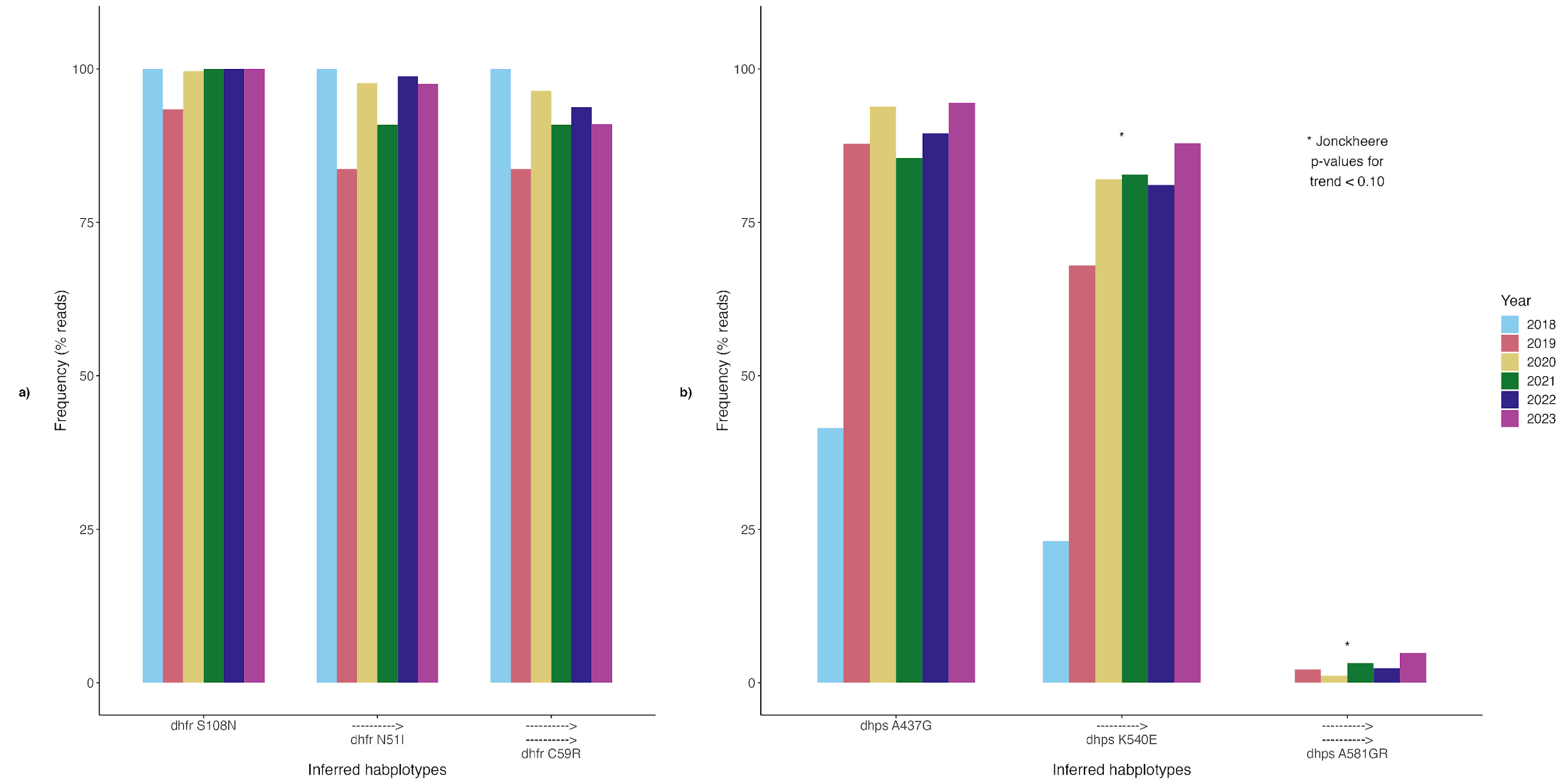

**Figure S3. Frequency of dhfr and dhps haplotypes associated with sulfadoxine-pyrimethamine.** Haplotypes were inferred from population level allele frequencies of markers of SP resistance. Jonckheere’s trend test was used to evaluate temporal changes in prevalence and frequency and an alpha value of 0.10 was used given the moderate sample size.

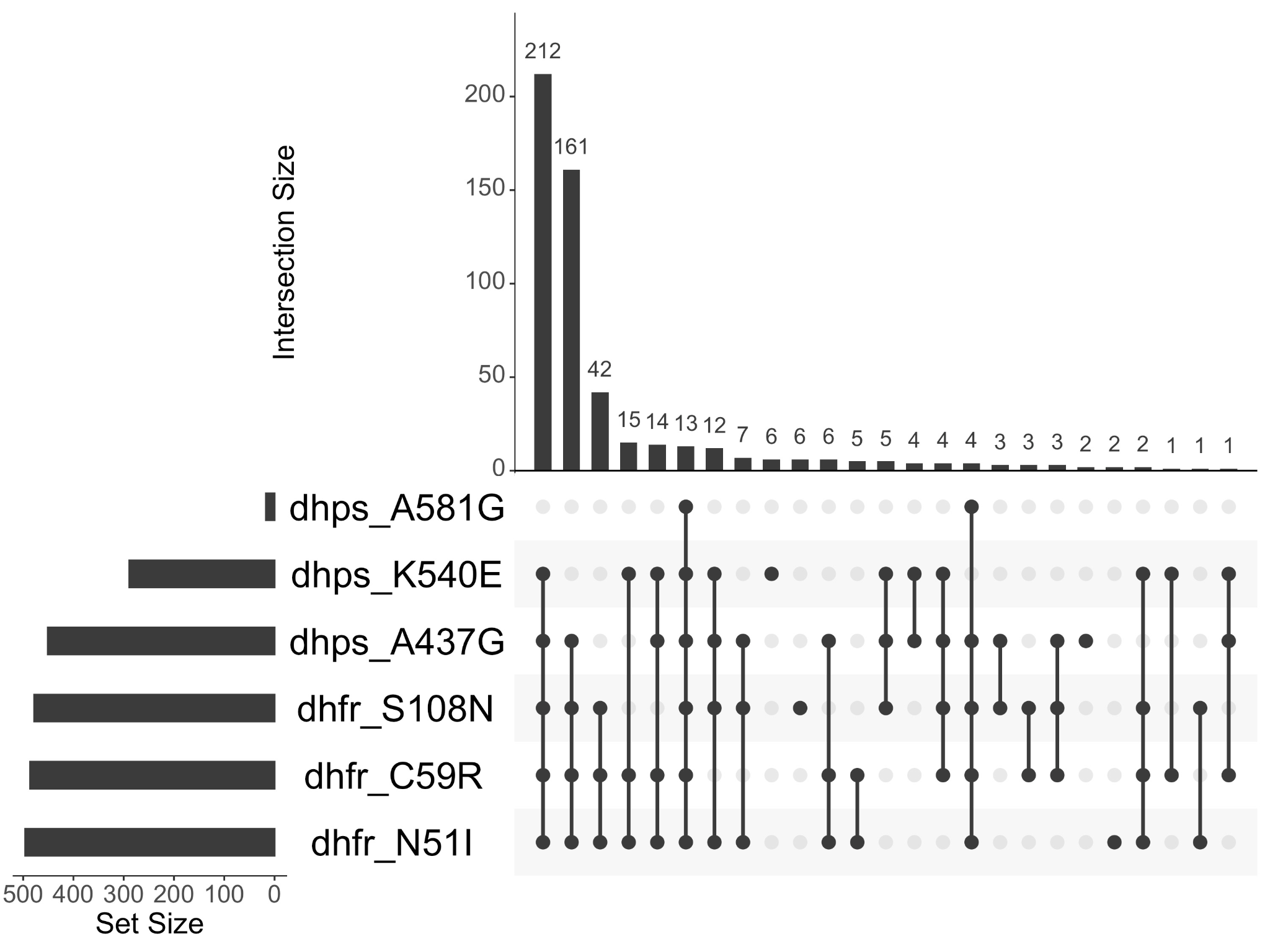

**Figure S4. Upset plot for SP resistance shows combinations of mutations found within the same sample.**

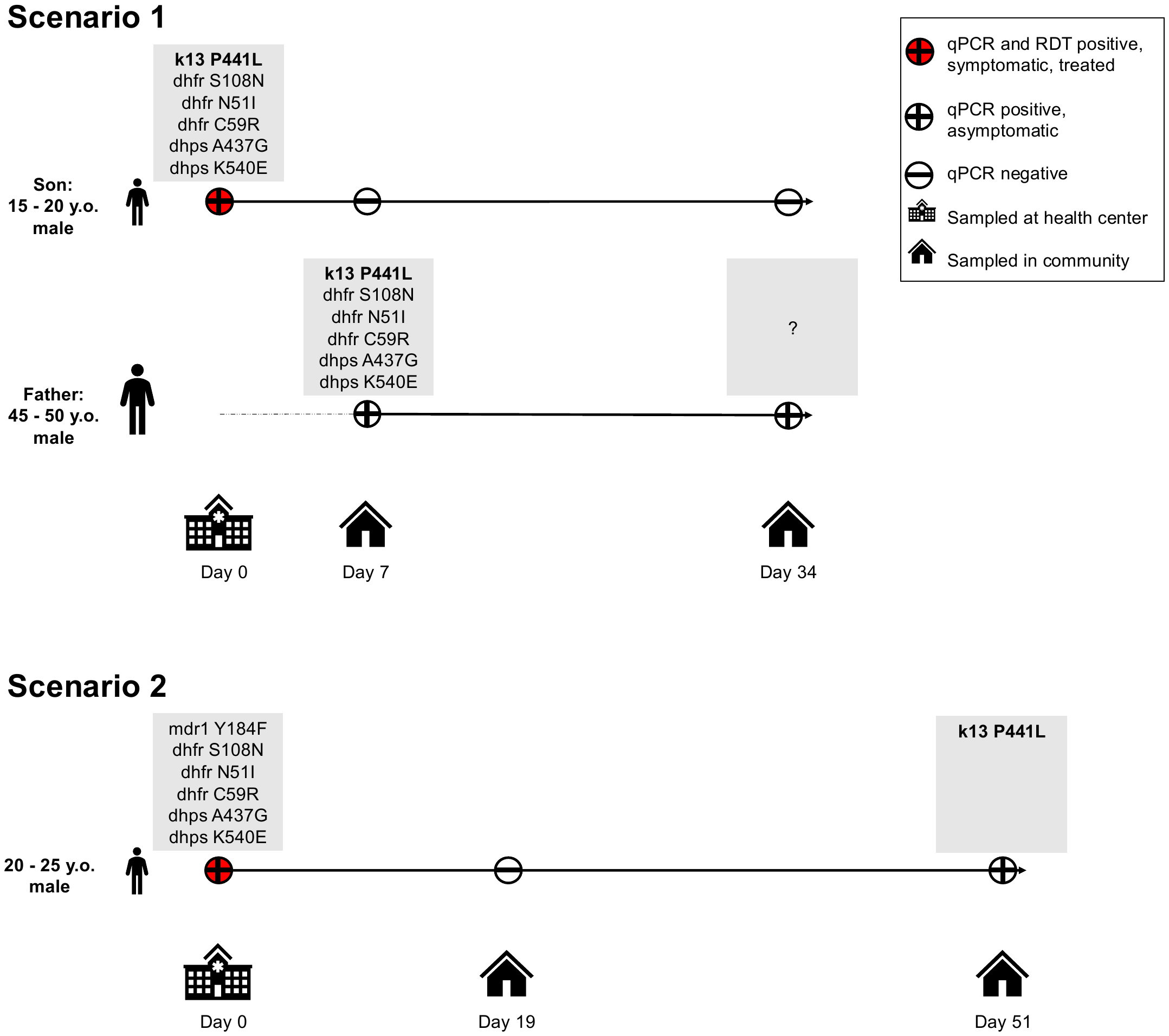

**Figure S5. Kelch mutations in community samples with epidemiological links to symptomatic samples.** Two asymptomatic cases with the P441L mutation identified through community surveillance were linked epidemiologically to samples containing drug resistant markers found in symptomatic individuals at the health center. In Scenario 1, a 15 – 20 year-old presented at the health center with symptomatic malaria, and his infection contained markers of sulfadoxine-pyrimethamine resistance and the P441L mutation. One week later, he and his family were sampled during community surveillance and his father, although asymptomatic, was positive for malaria by qPCR (parasite density of 3.4 parasites/µL), with the same set of mutations his son had earlier. One month later, the father was again asymptomatic but parasitemic, although his sample could not be amplified. Neither father nor son reported travel. In Scenario 2, a 20 – 25 year-old presented to the health center with symptomatic malaria and the parasites contained markers of sulfadoxine-pyrimethamine resistance and the mdr1 Y184F marker for multidrug resistance. He tested negative by qPCR at his community visit 19 days later but was again parasitemic (parasite density of 3 parasites/µL) 32 days later (Day 51 - Day 19). This sample contained the P441L mutation. Jonckheere’s trend test was used to evaluate temporal changes in prevalence and frequency and an alpha value of 0.10 was used given the moderate sample size (22).
